# Device-aided therapies (DATs) in Parkinson’s disease (PD). The DATs-PD GETM Spanish Registry Protocol Study

**DOI:** 10.1101/2024.12.07.24318643

**Authors:** D Santos-García, G González-Ortega, P Sánchez Alonso, A Planas-Ballvé, R García Ramos, I Cabo López, M Blázquez Estrada, A Sánchez Ferro, DATs-PD GETM Spanish Registry Group (Appendix 1)

**Author notes:** **Corresponding author:** Dr. Diego Santos García, Department of Neurology, Hospital Universitario de A Coruña (HUAC), Complejo Hospitalario Universitario de A Coruña (CHUAC), C/ As Xubias 84, 15006, A Coruña, Spain.

## Abstract

**Background and objective:** Device-aided therapies (DATs) are treatments indicated for people with Parkinsońs disease (PwP) with clinical fluctuations that are not optimally controlled with conventional medication. New DATs have recently emerged such as levodopa-entacapone-carbidopa intestinal gel infusion (LECIG) and subcutaneous infusion of foslevodopa/foscarbidopa (fLD/fCD). It is necessary to know the differences between different DATs.

**Patients and Methods:** We present here the protocol study of the DATs-PD GETM Spanish Registry. This is a descriptive, observational, prospective, multicenter, open study that is proposed as a clinical registry with progressive inclusion of PwP treated with a DAT in daily clinical practice conditions in more 40 centers from Spain for 10 years. The principal aim is to know the type of DAT that PwP in our country (Spain) receive. Specific objectives are to compare the clinical characteristics of the patients, the effectiveness, safety and tolerability, to identify predictors of a good response and to analyze the response by groups (gender, disease duration, phenotype, etc.). There is a baseline visit (V1; indication of the therapy), start visit (V2; initiation of the therapy) and follow-up visits at 6 months ± 3 months (V3_6M) and after this annually ± 3 months (V3_12M, V3_24M, etc.).

**Results:** The registry is on-going. The first patient was included on Abril 10, 2024. Patient recruitment and follow-up will be conducted until 31/DEC/2033. The estimate is to reach a minimum sample size of at least 3,000 patients.

**Conclusion:** The present study will help improve the care of PD patients treated with a DAT.

**Authors’ Roles:** **Santos-García** D: conception, organization, and execution of the project; funding acquisition; supervision; writing of the first draft of the manuscript; recruitment and/or evaluation of participants; entering data into the database. **González-Ortega** G: preparation and development of the database; registration of participants in the platform; recruitment and/or evaluation of participants; entering data into the database; review and critique. **Sánchez Alonso** P: recruitment and/or evaluation of participants; entering data into the database; review and critique. **Planas-Ballvé** A: recruitment and/or evaluation of participants; entering data into the database; review and critique. **García Ramos** R: recruitment and/or evaluation of participants; entering data into the database; review and critique. **Cabo López** I: recruitment and/or evaluation of participants; entering data into the database; review and critique. **Blázquez Estrada** M: recruitment and/or evaluation of participants; entering data into the database; review and critique. **Sánchez Ferro** A: collaboration in the coordination of the project as coordinator of GETM; recruitment and/or evaluation of participants; entering data into the database; review and critique.

**Financial Disclosures for the previous 12 months:** **Santos-García** D. has received honoraria for educational presentations and advice service by Abbvie, UCB Pharma, Lundbeck, KRKA, Zambon, Bial, Italfarmaco, Teva, Archímedes, Esteve, Stada, Merz, and grants from the “Fundación Professor Novoa Santos” as a result of the “CONVOCATORIA DE AYUDAS PARA LA REALIZACIÓN DE PROYECTOS DE INVESTIGACIÓN PARA GRUPOS EMERGENTES Y ASOCIADOS DEL INIBIC (2023/2024)”.

**González-Ortega** G. has received honoraria for educational purposes from ABBIE, Zambon, Bial, Esteve and Italfarmaco

**Sánchez Alonso** P. has received honoraria for educational presentations and advice service by Abbvie, UCB Pharma, Lundbeck, KRKA, Zambon, Bial, and Teva.

**Planas-Ballvé** A:None.

**García Ramos** R. has received honoraria and grants for lecturing, advisory services from Abbvie, Zambón, Bial, Merk, Stada.

**Cabo López** I. has received honoraria for educational presentations and advice service by Abbvie, Zambon, Bial, Orion, Italfarmaco and Esteve.

**Blázquez Estrada** M. has received honoraria for educational presentations by Dysport, Esteve, Bial, Italfármaco, Boston Sc. and Stada and for advice service by Esteve, Bial, Suazio.

**Sánchez Ferro** A. has received: grants or contracts from ERA-NET Horizon 2020 program JPCOFUND2 (reference number HESOCARE-329-073), MDS (eDiary project), Instituto de Salud Carlos III (reference number P122/01177); consulting fees from Abbvie, Esteve, Orion Pharma, and Prim; and payment or honoraria for lectures, presentations, speakers bureaus, manuscript writing, or educational events from Abbvie, Bayer, Esteve, MDS Society, EAN, Novartis, Monitor, Organon, Roche, SEN, Stada, Teva, and Zambon.

## INTRODUCTION

Parkinsońs disease (PD) is the second most common neurodegenerative disease after Alzheimer’s disease. It is characterized by a deficit of dopamine in the striatum and other brain areas, but also of other neurotransmitters such as noradrenaline, acetylcholine or serotonin, which would explain the appearance of motor and non-motor symptoms characteristic of the disease [1]. The diagnosis is made by applying well-defined criteria [2,3] that are based fundamentally on the existence of parkinsonism, the absence of atypical data that suggest an alternative diagnosis (pharmacological, Parkinson-Plus syndrome, etc.), and data in favor that suggest PD itself. Among the latter, a determining aspect is the good response to dopaminergic medication, especially levodopa [4)]. Its administration compensates for the deficit of dopaminergic stimulation at the level of the postsynaptic receptors, which causes an improvement in symptoms such as tremor, rigidity or bradykinesia, among others. In fact, the absence of a response would go against the diagnosis of PD.

Although the response to dopaminergic medication may be optimal during the first few years, people with PD (PwP) develop motor and non-motor complications (e.g., motor fluctuations, non-motor fluctuations, dyskinesias, etc.) that impact their quality of life and autonomy [5]. Thus, PwP may only perceive improvement at certain times of the day alternating with disabling OFF episodes where the symptoms reemerge and the control with conventional medication is insufficient [6]. Some PwP may benefit from treatment with a second-line therapy that is an alternative and/or complement to conventional medication that is not sufficient [7]. The proper selection of the candidates is key, and there are different tools for their correct identification [8]. These therapies are commonly known as device-aided therapies (DATs) and although they are more expensive and complex than conventional medication, they have been shown to reduce OFF time, improve non-motor symptoms and the quality of life of PwP [9]. For many years the available DATs in many countries have been the deep brain stimulation (DBS), continuous subcutaneous apomorphine infusion (CSAI) and levodopa-carbidopa intestinal gel infusion (LCIG) [10]. However, new treatments have recently emerged, such as levodopa-entapacone-carbidopa intestinal gel infusion (LECIG) and subcutaneous infusion of foslevodopa-foscarbidopa (fLD/fCD) [11,12]. Particularly relevant is the recent availability of fLD/fCD subcutaneous infusion to the point that many recently published treatment algorithms are outdated and the possibility of considering the subcutaneous route as the first alternative to the enteral route is being discussed, as it is less invasive when considering infusion therapy [13,14].

Therefore, a completely new scenario is opening up in the treatment of PwP with a DAT and it is essential to know the frequency of the prescriptions, the characteristics of the treated individuals as well as their long-term evolution in relation to clinical changes and complications. This is the reason why, from the Movement Disorders Study Group (Grupo de Estudio de Trastornos de Movimiento [GETM]) of the Spanish Neurological Society (Sociedad Española de Neurología [SEN]), we have launched a prospective registry of PwP treated with a DAT in our country (Spain). The aim of this article is to describe this registry that we have named DATs-PD GETM Spanish Registry.

## METHODS/DESIGN

### Title of the Project

Device-aided therapies in Parkinsońs disease GETM Spanish Registry (DATs-PD GETM Spanish Registry).

### Type of study

Descriptive, observational, prospective, multicenter, open study that is proposed as a clinical registry with progressive inclusion of patients with PwP treated with a DAT in daily clinical practice conditions.

### Promoter and principal coordinator of the project

Diego Santos García, MD, PhD.

### Coordinating institutions

“Fundación Degen”, “Grupo de Estudio de Trastornos del Movimiento (GETM)” and “Sociedad Española de Neurología (SEN)”.

### Participating centers in the project

More than 40 centers from Spain with neurology teams with experience in the management of PD (**Appendix 1**), including the use of DATs.

### Population

PwP treated with a DAT in Spain from year 2024. The following therapies are included as DATs: DBS; CSAI; LCIG; LECIG; and fLD/fCD subcutaneous infusion. In addition, PwP who are treated with focused ultrasound-guided thermal ablation will also be included. Specifically, the eligibility criteria are: 1) diagnosis of PD according to the MDS criteria; 2) start of treatment with a DAT from January 1, 2024; 3) the patient’s desire to participate on a completely voluntary basis; 4) signing of an informed consent.

### Justification of the project

1) there are many DATs currently used to treat PwP; 2) some of them have been introduced very recently and we have no experience; 3) in this context, we must know about the indication preferences in Spanish centers that treat PwP; 4) we must know the characteristics of the individuals treated and the differences between therapies; 4) we must know the effectiveness of DATs and to compare them; 5) we must know the safety and tolerability of DATs and to compare them; 6) it is necessary to define different response profiles or predictors of better or worse outcome; 7) this is an excellent opportunity to be able to develop a registry at this time that collects a lot of information over the long term; 8) It could be a starting point to which centers from other countries can join and develop a broader registry (e.g., European, etc.).

## Objectives

### Principal objective

to know the type of DAT that PwP in our country receive, treated by expert neurologists in daily clinical practice conditions.

### Specific objectives

1) to analyze the sociodemographic and clinical characteristics of PwP treated with a DAT, comparing the different treatments (DBS vs subcutaneous vs enteral treatment); 2) to analyze the effectiveness of different DATs, comparing the different treatments (DBS vs subcutaneous vs enteral treatment); 3) to analyze the safety and tolerability of different DATs, comparing the different treatments; 4) specifically, to analyze the changes experienced by PwP treated with DATs in the perception of their quality of life and autonomy for carrying out daily activities and to compare the different treatments; 5) specifically, to compare the rate of maintenance of the therapy between the different DAT groups; 6) specifically, to compare the dropout rate for each of the therapies and find out the underlying reasons; 7) to find out the reasons for changing from one therapy to another or adding a second DAT to a previous one being received; 8) to analyze the complications in relation to the different DATs and compare them; 9) to compare the differences by gender (male vs. female); 10) others that may be raised based on the data collected (e.g., differences by age, disease duration etc.).

### Visits

The registry includes 3 types of visits: 1) baseline visit (V1), which is when the DAT is decided by the neurologist; 2) start visit (V3), which is when the DAT is initiated by the patient; 3) follow-up visit (V3), with the patient receiving the DAT. The first follow-up visit will be carried out at 6 months +/-3 months (V3_6M) and then at 1 year +/-3 months and subsequently annually +/-3 months: 1 year (V3_12M); 2 years (V3_24M); 3 years (V3_36M); 4 years (V3_48M); 5 years (V3_60M); etc. (**Figure 1**).

**Figure 1.**
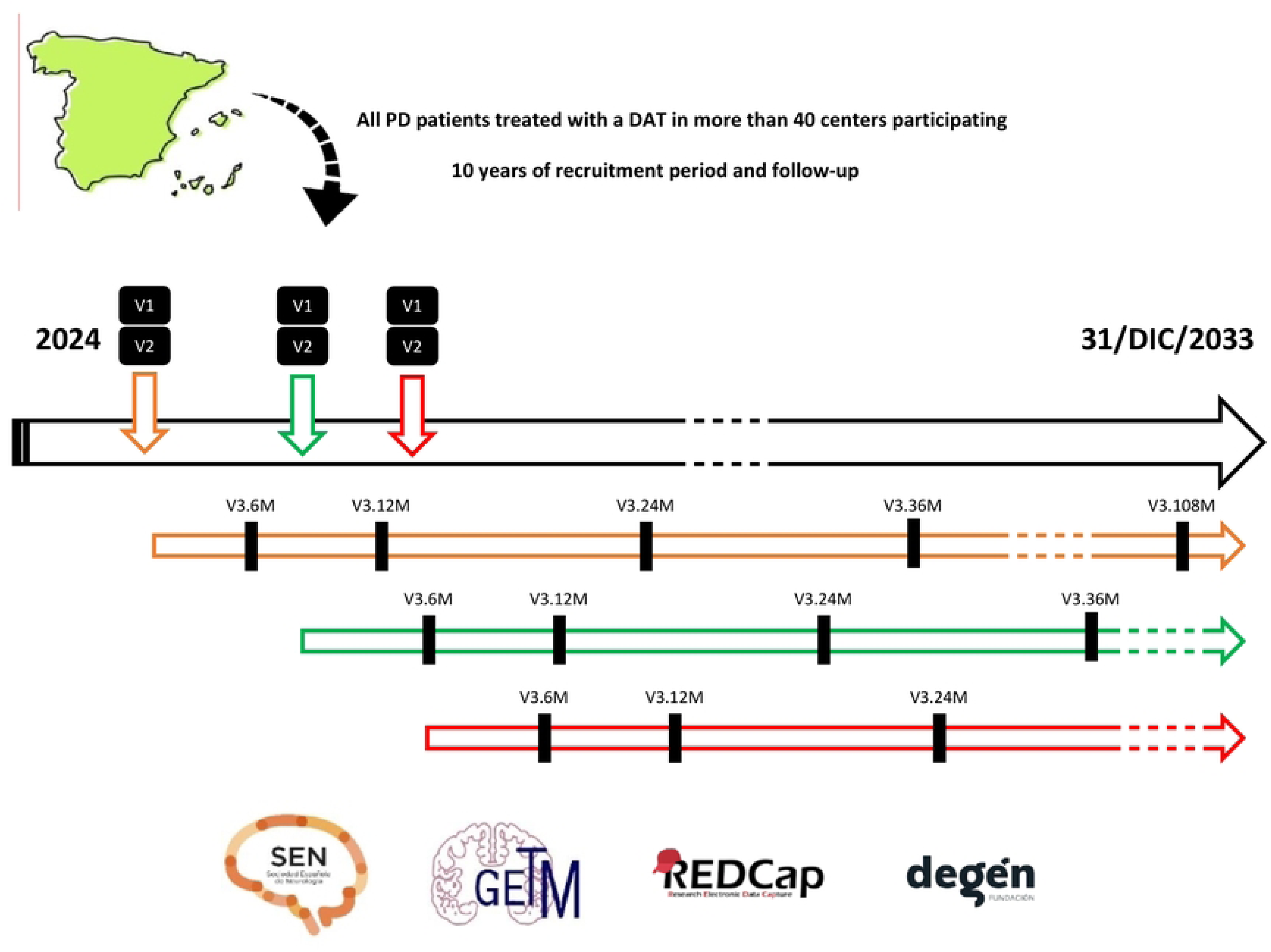
All patients treated with a DAT from 2024 to 31/DEC/2033 in more than 40 participating centers from Spain are included, all of them evaluated by neurologists who are experts in PD. Each color refers to a different patient. There is a baseline visit (V1; indication of the therapy), start visit (V2; initiation of the therapy) and follow-up visits at 6 months (V3_6M) and after this annually (V3_12M, V3_24M, etc.). The window for V2 and V3 is ± 3 months.

The period of inclusion of patients in the registry will be a minimum of 5 years, extendable to 10 years according to the approved protocol. New PwP will be included over a 10-year period, while each participant’s included will be follow-up during this time. The data collection period ranges from January 1, 2024 to December 31, 2033. If a person with PD switches to another DAT or a second DAT is added, the information will be collected. In addition, if a subject drops out of the DAT and does not receive another DAT, data collection will continue within the registry to allow for a comparative arm of untreated patients after a therapeutic failure (something rarely discussed in the literature). Although the registry is prospective, it is planned to include PwP treated in 2024 whose indication for DAT was before this year, as long as the data was properly collected in the medical record. An example is DBS, given that the waiting list from indication to intervention is long in some centers.

### Assessments

The following information will be collected at each visit:

1. Baseline visit (V1): sociodemographic data; data about PD (age onset, disease duration, motor phenotype, etc.); comorbidities; treatments; main reason for therapy indication; levodopa equivalent daily dose (LEDD) [15]; motor symptoms; non-motor symptoms; Unified Parkinsońs Disease Rating Scale (UPDRS-III) [16] during the OFF state; UPDRS-III during the ON state; Hoehn & Yahr (H&Y) [17] stage in OFF; H&Y stage in ON; MNCD classification (classification; score; stage) [18]; Parkinson’s Disease Questionnaire (PDQ-39) [19]; EUROHIS-QOL 8-item index [20]; Schwab and England Activities of Daily Living Scale (ADLS) [21] during the OFF and during the ON state; serum levels of folate, B vitamins (B1, B6, B12) and homocysteine.
2. Start visit (V2): data about the DAT and treatment settings including LEDD.
3. Follow-up visit (V3): data about the DAT and treatment settings including LEDD; comorbidities; treatments; LEDD; motor symptoms; non-motor symptoms; UPDRS-III during the ON state; H&Y stage in ON; MNCD classification (classification; score; stage); Parkinson’s Disease Questionnaire (PDQ-39); EUROHIS-QOL 8-item index; ADLS during the OFF and during the ON state; serum levels of folate, B vitamins (B1, B6, B12) and homocysteine; complications about the DAT and/or other relevant complications.

All information will be collected by expert neurologists in a daily clinical practice setting. A total of 16 and 26 questions collect information on motor symptoms/complications and non-motor symptoms, respectively, which are categorized as absent, mild, moderate, severe, or very severe. The score on the MDS-UPDRS-III scale [22] will be an added option that will be collected in those centers that use it (permission by the MDS was obtained). **Table 1** summarizes the information on the variables that will be collected during the different visits.

**Table 1.** Variables collected in the DAT-PD Spanish Registry according to the type of visit.

| V1 | V2 | V3 |
| --- | --- | --- |
| Sociodemographic<br>PD history<br>Comorbidities<br>Concomitant treatments<br>LEDD<br>Main reason for DAT<br>PD symptoms<br>Motor symptoms<br>Non-motor symptoms<br>Motor scales<br>UPDRS-III-OFF<br>UPDRS-III ON<br>H&Y-OFF<br>H&Y-ON<br>Quality of life<br>PDQ-39<br>EUROHIS-QOL8<br>Functional dependency<br>ADLS-OFF<br>ADLS-ON<br>Serum biomarkers<br>Folate<br>Vitamin B1<br>Vitamin B6<br>Vitamin B12<br>Homocysteine | Information about DAT<br>Concomitant treatments<br>LEDD | Information about DAT<br>Concomitant treatments<br>LEDD<br>PD symptoms<br>Motor symptoms<br>Non-motor symptoms<br>Motor scales<br>UPDRS-III-OFF<br>UPDRS-III ON<br>H&Y-OFF<br>H&Y-ON<br>Quality of life<br>PDQ-39<br>EUROHIS-QOL8<br>Functional dependency<br>ADLS-OFF<br>ADLS-ON<br>Serum biomarkers<br>Folate<br>Vitamin B1<br>Vitamin B6<br>Vitamin B12<br>Homocysteine<br>Complications<br>Related with DAT<br>Unrelated with DAT |

ADLS, Schwab & England Activities of Daily Living Scale; DAT, device-aided therapy; LEDD, levodopa equivalent
daily dose; PD, Parkinson’s disease; PDQ-39, the 39-item Parkinson’s disease Questionnaire; UPDRS, Unified
Parkinson’s Disease Rating Scale.

### Data collection and statistical analysis

Data will be collected using REDCap. This is a secure web application for building and managing online surveys and databases. REDCap has been used to date (27/NOV/2024) in 159 countries by more than 3.4 million users. Data collected will be transferred to a statistical package for subsequent analysis. The promoter team of the project will be responsible for study monitoring. The estimate, with the participation of more than 40 centers and a recruitment period of 10 years, is to be able to reach a minimum sample size of 3,000 patients. The pertinent analysis (descriptive, missing data analysis, normality assumptions, univariate, binary logistic regression, multiple linear regression, etc.) will be performed based on the type of objective. In addition, given de complexity of potential analysis including a diversity of variables from different origin and measurement properties, advanced statistical methodology (data mining, machine learning or other artificial intelligence techniques, etc.) could be considered when applicable.

### Standard protocol approvals, registrations, and patient consents

The project will be conducted in accordance with the ICH Good Clinical Practice version 6 Revision 2 standard, the fundamental ethical principles established in the Declaration of Helsinki and the Oviedo Convention, as well as the Spanish legal requirements for biomedical research (Biomedical Research Law 14/2007). The Project has been approved on 02/APR/2024 by the IRB “Comité de Ética de la Investigación Clínica de Galicia from Spain” with code number 2024/109. Written informed consents from all participants in this study will be obtained.

### Data availability

The protocol, statistical analysis plan and deidentified participant data will be available on request.

### Study timetable

1. Pre-start-up procedures: until April 2024.
2. First patient included: 10/APR/2024.
3. First analysis to present: 2025 – Q1 (data about PD patients treated in 2024).
4. Proposed specific objectives and others: from 2025 onwards.

### Funding statement

An amount of 25.000 euros has been granted by the “Fundación Professor Novoa Santos” as a result of the “CONVOCATORIA DE AYUDAS PARA LA REALIZACIÓN DE PROYECTOS DE INVESTIGACIÓN PARA GRUPOS EMERGENTES Y ASOCIADOS DEL INIBIC (2023/2024)”. The import will be used for data monitoring and other actions required in the context of the project.

### Strengths and limitations of the project

The main strength of the project is the possibility of collecting systematic information on patients treated with DATs over a long follow up period (10 years). Further, there is great interest in the fact that some of these therapies are novel and the management of advanced PD patients is changing because of this and could hence be evaluated. Also, the methodology with prospective follow-up and the collection of well-defined variables (UPDRS, H&Y, PDQ-39, ADLS, etc.) in a clinical practice setting will allow generalizing the findings to other environments. To this end, we also consider in the future increasing the number of centers in Spain, and even the chance of extrapolating the registry to other countries harmonizing the data collected. Other future possibilities are implementing new technologies to monitor the disease and analyze the changes after starting with the DAT and their correlation with the data collected from the registry.

On the other hand and as limitations, unlike a study with a clearly defined protocol in which multiple scales are administered [23], this project is a registry, and limitations in the quantity and quality of the data collected are possible. For example, the number and description of complications may be underestimated. At this moment the project is exciting but there is some residual risk that it will not work properly due to the workload of researchers.

## DISCUSSION

According to the PARADISE study, a non-interventional, cross-sectional, multicenter, national study conducted in the hospital setting and published in 2021, up to 38.2% of people with PD in Spain have advanced PD [24]. The key aspect of identifying advanced PD is to differentiate which PwP could be candidates to receive a DAT. However, only 15.2% of PwP from the advanced PD group from the PARADISE study were receiving some form of therapy for advanced stages of the disease (i.e., DBS, CSAI, LCIG). The most frequent reasons why advanced PwP were not on a DAT were “to be clinically stable” and “option not yet considered”. Similar results were observed in the PROSPECT study [25], in which only 20.9% of PwP (adults with levodopa-responsive PD and inadequately controlled motor symptoms with ≥ 2.5-hours/day “Off” time, despite trials of available oral/transdermal/sublingual/inhalable medication) initiated a DAT despite the fact that, as expected, DAT vs best medical therapy improved motor symptoms, non-motor symptoms, sleep, quality of life, independence for ADL and caregiver burden. For many years, the options for being treated with a DAT meant choosing between three alternatives: DBS, CSAI, LCIG. However, the scenario is new with the current availability of LECIG and subcutaneous fLD/fCD. In particular, it is of maximum interest to know whether the arrival of fLD/fCD implies a reduction in the indication of other treatments and what are the characteristics of the treated patients as well as their complications in comparison with other DATs, especially enteral therapies [26]. This registry aims to answer this question and many others explained in the objectives. We believe that the information collected in the DATs-PD GETM Spanish Registry will be of great interest in helping to improve the management of people with Parkinson’s disease treated with a DAT.

In summary, we present here the protocol study of the DATs-PD GETM Spanish Registry, a descriptive, observational, prospective, multicenter, open study that is proposed as a clinical registry with progressive inclusion of PwP treated with a DAT in daily clinical practice conditions in more 40 centers from Spain for 10 years. The present study will help improve the care of PwP treated with a DAT.

## Data Availability

No results are shown. This is a Study Protocol. The protocol, statistical analysis plan and deidentified participant data will be available on request (for this project, the registry).

## Abbreviations

ADL: activities of daily living
CSAI: continuous subcutaneous apomorphine infusion
DATs: device-aided therapies
DBS: deep brain stimulation
fLD/fCD: foslevodopa-foscarbidopa
LCIG: levodopa-carbidopa intestinal gel infusion
LECIG: levodopa-entacapone-carbidopa intestinal gel infusion
LEED: levodopa equivalent daily dose
PD: Parkinsońs disease
UPDRS: Unified Parkinsońs Disease Rating Scale.

## Acknowledgements

We would like to thank all patients who collaborated in this study. Many thanks also to Fundación Española de Ayuda a la Investigación en Enfermedades Neurodegenerativas y/o de Origen Genético (https://fundaciondegen.org/), Grupo de Estudio de Trastornos del Movimiento (GETM), Sociedad Española de Neurología (SEN) and Fundación Profesor Novoa Santos.

## Appendix 1 DATs-PD GETM Spanish Registry STUDY GROUP

Adarmes Gómez AD, Alonso Modino D, Álvarez Sauco M, Aneiros Díaz A, Ávila Rivera A, Baviera-Muñoz R, Belmonte S, Blázquez Estrada M, Caballero Sánchez L, Caballol Pons N, Cabo López I, Campins Romeu M, Campolongo A, Cantarero Duque S, Carrillo García F, Casanova Mollá J, Casas Peña E, Castaño García B, Castellano Guerrero AM, Castrillo Sanz A, Cerdán Santacruz DM, Clavero Ibarra P, Cores Bartolomé C, Cots Foraster A, Cubo Delgado E, Delgado Ballestero T, Erdocia Goñi A, Escalante Arroyo S, Escamilla Sevilla F, Espinosa Rosso R, Fanjul Arbos S, Feliz Feliz C, Fernández Pajarín G, Fernández Revuelta A, Fernández Rodríguez B, Fernández Valle T, Freire Álvarez E, Gamo González E, García Fernández C, García Herruzo A, García Ramos García R, García Ruíz Espiga P, Garrote Espina L, Gil Villar MP, Gómez Esteban JC, Gómez López de San Román C, Gómez Mayordomo V, Gómez Rapela C, González MV, González Ardura J, González Hernández A, González-Ortega G, Guerra Hiraldo JD, Gutiérrez García J, Hernández Vara J, Jesús Maestre S, Kulisevsky Bojarski J, Legarda Ramírez I, López Ariztegui N, López Dominguez D, López Manzanares L, López Veloso AC, López Valdés E, Lorenzo Barreto P, Lorenzo Brito JM, Lozano D, Macías García D, Madrid Navarro CJ, Martí Andrés G, Martí Martínez S, Martín García R, Martínez Castrillo JC, Martínez-Torres I, Mata Álvarez Santullano M, Mauri Fabrega L, Méndez Guerrero A, Mendoza Rodríguez A, Mir Rivera P, Mondragón Rezola E, Monterde Ortega A, Morales Casado MI, Morata-Martínez C, Muñoz Delgado L, Muñoz Ruíz T, Muro I, Novo Ponte S, Ojeda Lepe E, Olivares Romero J, Pagonabarraga J, Pareés Moreno I, Pascual Sedano B, Paz González JM, Peral Quirós A, Pérez Calvo C, Pérez Rangel D, Perona Moratalla A, Planas-Ballvé A, Prendes Fernández P, Quibus Requena L, Rábano Suárez P, Rashid López R, Rebollo Lavado B, Rojas Pérez E, Ribacoba Díaz C, Romero Fábrega JC, Ruiz López M, Ruíz Martínez J, Samaniego Vinueza LB, San Eufrasio Martínez M, Sánchez Alonso P, Sánchez Ferro A, Sánchez Rodríguez A, Sancho Saldana A, Santos-García D, Sastre Bataller I, Sesar Ignacio A, Solano Vila B, Solleiro Vidal A, Suárez San Martín E, Tabar Comellas G, Tijero Merino B, Triguero Cueva L, Valero García MF, Valldeoriola Serra F, Vela L, Vinagre Aragón A, Vivas Villacampa L, Vives Pastor B, Yáñez Baña R.

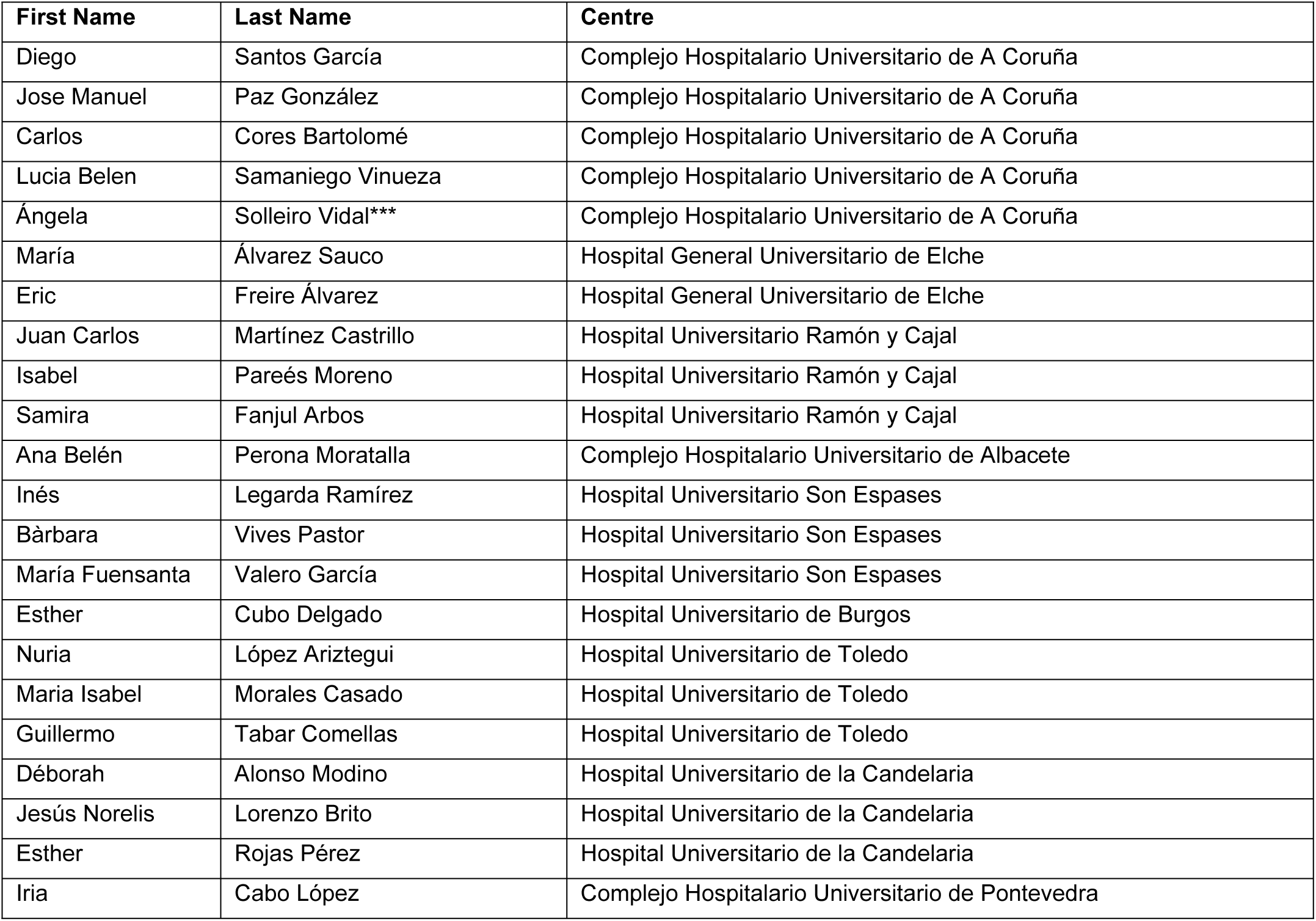
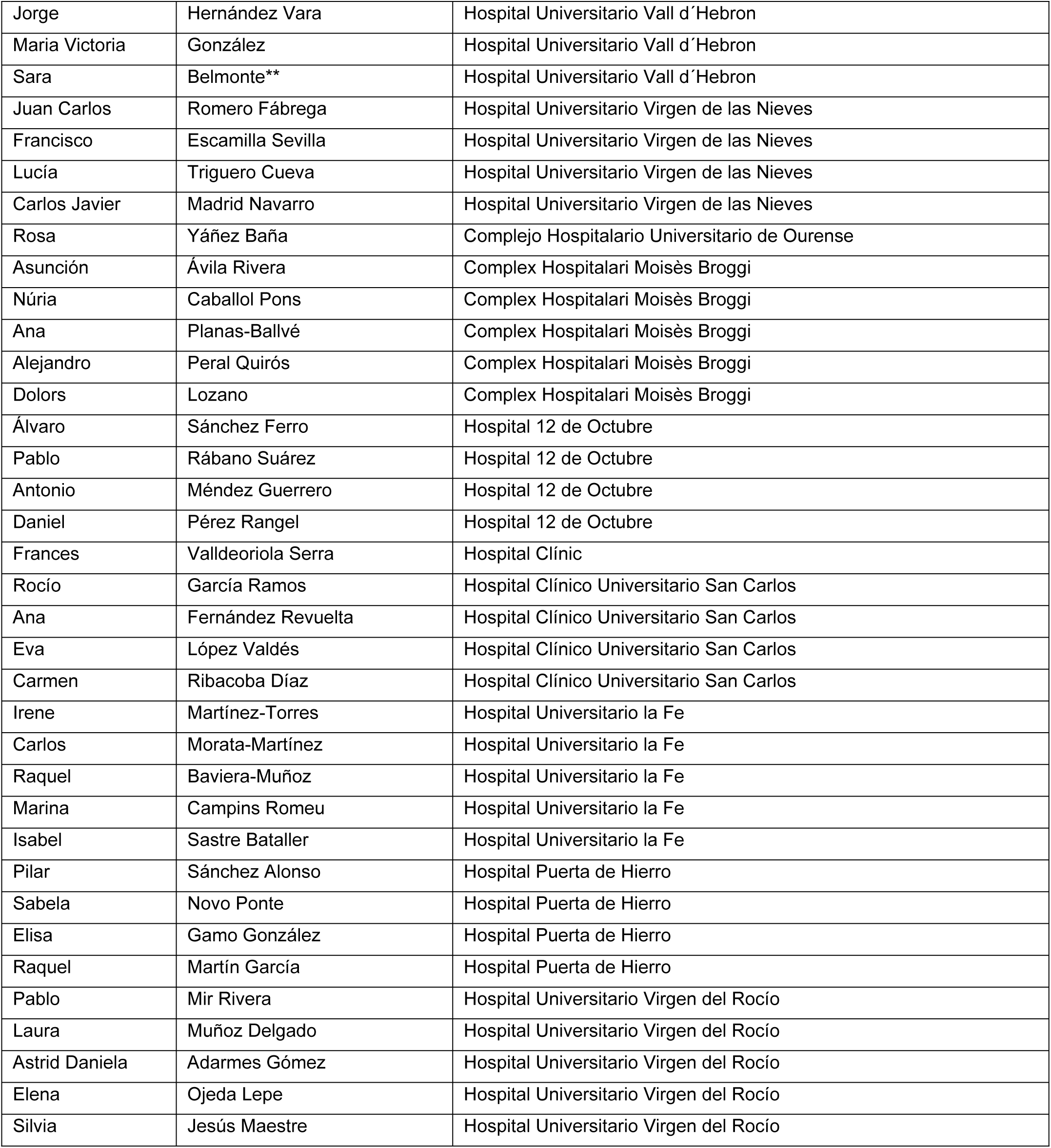
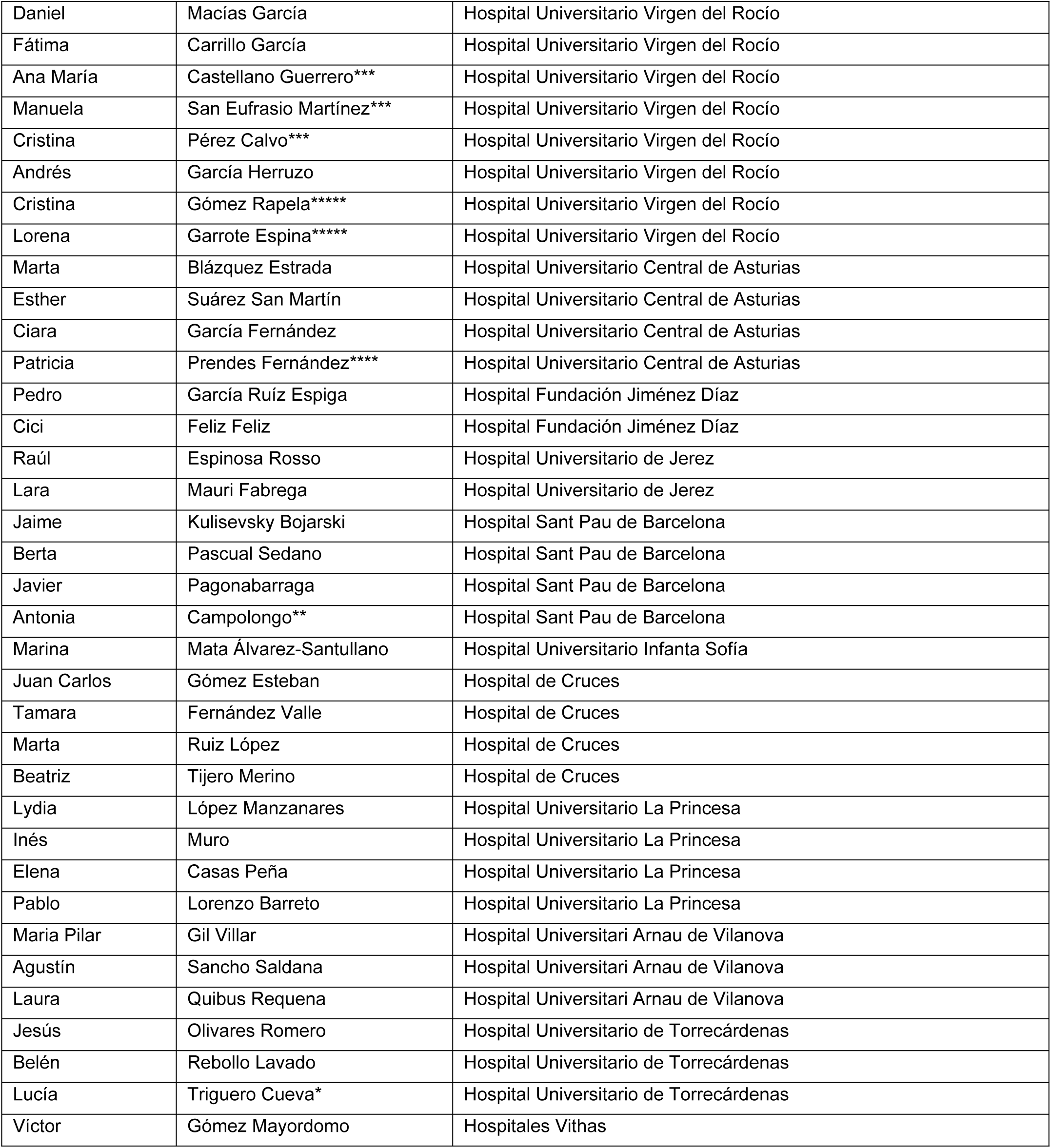
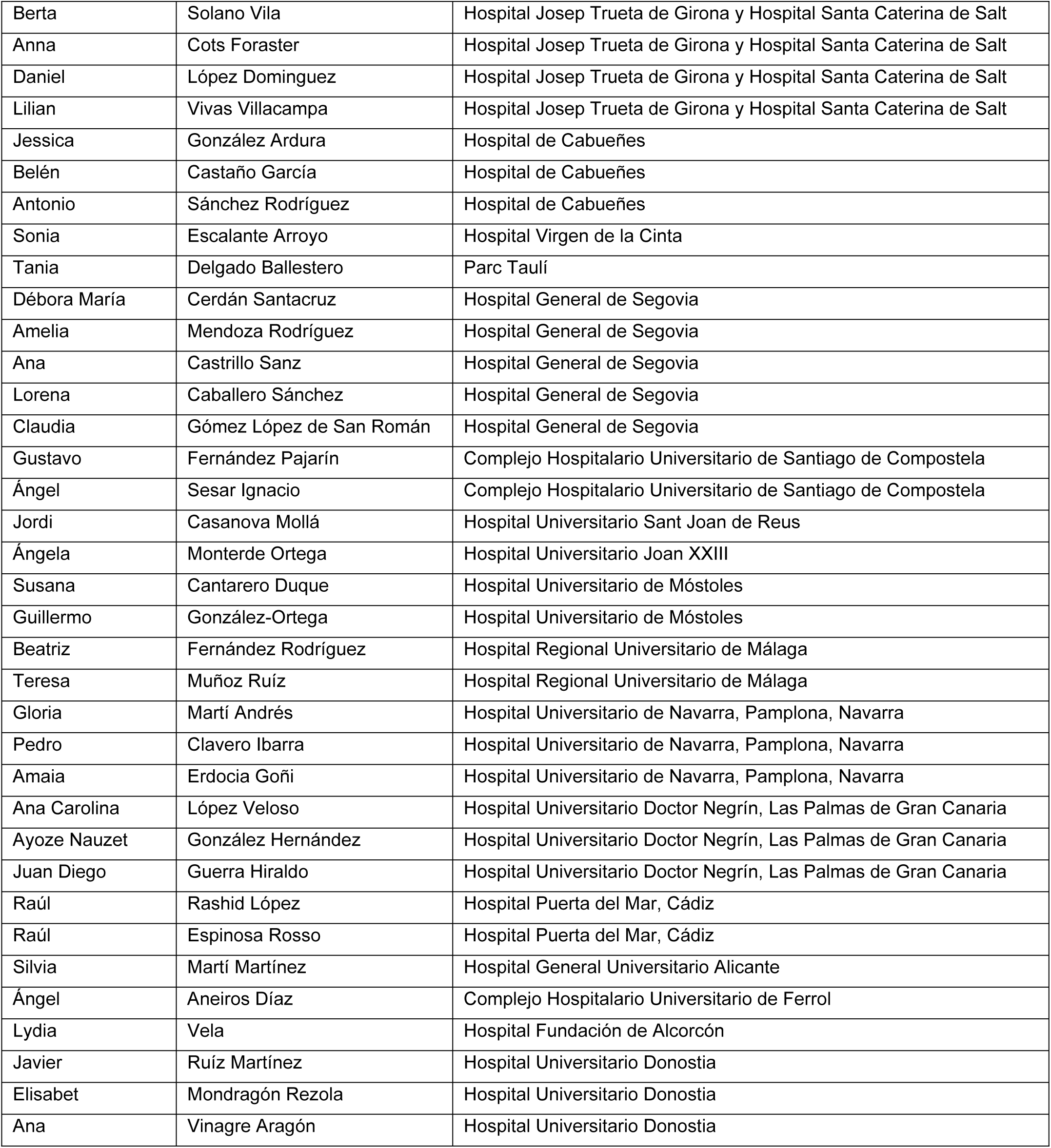
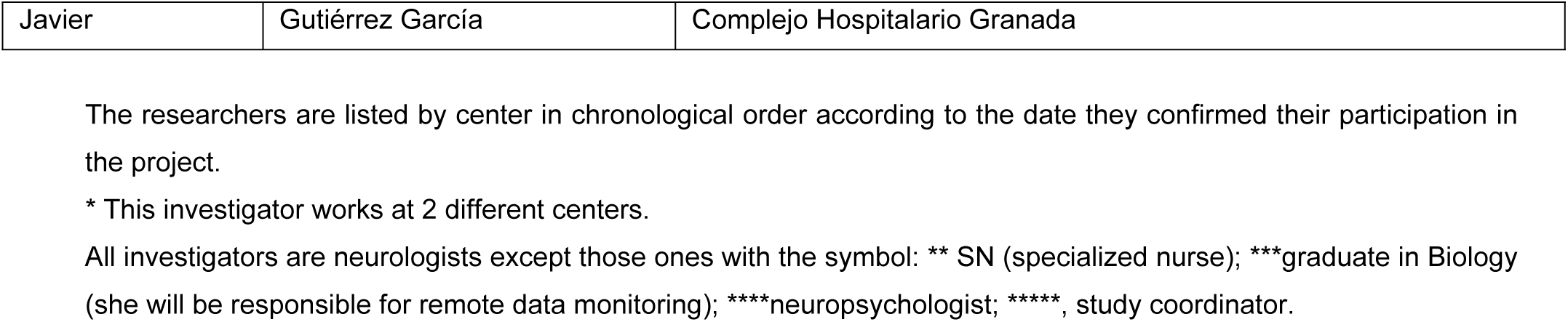

## Notes

### Competing Interest Statement

The authors have declared that no competing interests exist.

### Funding Statement

The author(s) received no specific funding for this work.

### Author Declarations

The Project has been approved on 02/APR/2024 by the IRB “Comité de Ética de la Investigación Clínica de Galicia from Spain” with code number 2024/109. Written informed consents from all participants in this study will be obtained.

## References

1. Bloem BR, Okun MS, Klein C. Parkinson’s disease. Lancet 2021;397:2284–303.

2. Hughes AJ, Daniel SE, Kilford L, Lees AJ. Accuracy of clinical diagnosis of idiopathic Parkinson’s disease: a clinico-pathological study of 100 cases. J Neurol Neurosurg Psychiatry 1992;55:181–4.

3. Postuma RB, Berg D, Stern M, Poewe W, Olanow CW, Oertel W, et al. MDS clinical diagnostic criteria for Parkinson’s disease. Mov Disord 2015;30:1591–601.

4. Santos-García D, de us Fonticoba T, Cores Bartolomé C, et al.; COPPADIS Study Group. Response to levodopa in Parkinson’s disease over time. A 4-year follow-up study. Parkinsonism Relat Disord 2023;116:105852.

5. Hauser RA, McDermott MP, Messing S. Factors associated with the development of motor fluctuations and dyskinesias in Parkinson disease. Arch Neurol 2006;63:1756–60.

6. Kakimoto A, Kawazoe M, Kurihara K, Mishima T, Tsuboi Y. Impact of non-motor fluctuations on QOL in patients with Parkinson’s disease. Front Neurol 2023;14:1149615.

7. Deuschl G, Antonini A, Costa J, et al. European Academy of Neurology/Movement Disorder Society-European Section Guideline on the Treatment of Parkinson’s Disease: I. Invasive Therapies. Mov Disord 2022;37:1360–74.

8. Moes HR, Henriksen T, Sławek J, Phokaewvarangkul O, Buskens E, van Laar T. Tools and criteria to select patients with advanced Parkinson’s disease for device-aided therapies: a narrative review. J Neural Transm (Vienna) 2023;130:1359–77.

9. Phokaewvarangkul O, Auffret M, Groppa S, Markovic V, Petrovic I, Bhidayasiri R. What was first and what is next in selecting device-aided therapy in Parkinson’s disease? Balancing evidence and experience. J Neural Transm (Vienna) 2024;131:1307–20.

10. Auffret M, Weiss D, Stocchi F, Vérin M, Jost WH. Access to device-aided therapies in advanced Parkinson’s disease: navigating clinician biases, patient preference, and prognostic uncertainty. J Neural Transm (Vienna) 2023;130:1411–32.

11. Santos-García D, López-Manzanares L, Muro I, at el. Effectiveness and safety of levodopa-entacapone-carbidopa infusion in Parkinson disease: A real-world data study. Eur J Neurol 2024 (epub ahead of print).

12. Soileau MJ, Aldred J, Budur K, et al. Safety and efficacy of continuous subcutaneous foslevodopa-foscarbidopa in patients with advanced Parkinson’s disease: a randomised, double-blind, active-controlled, phase 3 trial. Lancet Neurol 2022;21(12):1099–109.

13. Antonini A, D’Onofrio V, Guerra A. Current and novel infusion therapies for patients with Parkinson’s disease. J Neural Transm (Vienna) 2023;130:1349–58.

14. Aubignat M, Tir M. Continuous Subcutaneous Foslevodopa-Foscarbidopa in Parkinson’s Disease: A Mini-Review of Current Scope and Future Outlook. Mov Disord Clin Pract 2024;11:1188–94.

15. Nyholm D, Jost WH. An updated calculator for determining levodopa-equivalent dose. Neurol Res Pract 2021;3:58.

16. Fanhn S, Elton RL, Members of the UPDRS Development Committee. Unified Parkinson’s Disease Rating Scale. In: Fahn S, Marsden CD, Calne DB, Goldstein M, editors. Recent developments in Parkinson’s disease, Vol 2. Florham Park, NJ: Macmillan Health Care Information;1987.p-153–64.

17. Hoehn MM, Yahr MD. Parkinsonism: onset, progression and mortality. Neurology 1967;17:427–42.

18. Santos García D, Álvarez Sauco M, Calopa M, et al. MNCD: A New Tool for Classifying Parkinson’s Disease in Daily Clinical Practice. Diagnostics (Basel) 2021;12:55.

19. Jenkinson C, Fitzpatrick R, Peto V, Greenhall R, Hyman N. The Parkinson’s Disease Questionnaire (PDQ-39): development and validation of a Parkinson’s disease summary index score. Age Ageing 1997;26:353–7.

20. Da Rocha NS, Power MJ, Bushnell DM, Fleck MP. The EUROHIS-QOL 8-item index: comparative psychometric properties to its parent WHOQOL-BREF. Value Health 2012;15:449–57.

21. Schwab RS, England AC. In Third Symposium on Parkinson’s disease. E. And S. Livingstone: Edinburgh 1969;152-7.

22. Goetz CG, Tilley BC, Shaftman SR, et al.; Movement Disorder Society UPDRS Revision Task Force. Movement Disorder Society-sponsored revision of the Unified Parkinson’s Disease Rating Scale (MDS-UPDRS): scale presentation and clinimetric testing results. Mov Disord 2008;23:2129–70.

23. Santos-García D, Mir P, Cubo E, Vela L, et al.; COPPADIS Study Group. COPPADIS-2015 (COhort of Patients with PArkinson’s DIsease in Spain, 2015), a global--clinical evaluations, serum biomarkers, genetic studies and neuroimaging--prospective, multicenter, non-interventional, long-term study on Parkinson’s disease progression. BMC Neurol 2016;16:26.

24. Martínez-Castrillo JC, Martínez-Martín P, Burgos Á, et al. Prevalence of Advanced Parkinson’s Disease in Patients Treated in the Hospitals of the Spanish National Healthcare System: The PARADISE Study. Brain Sci 2021;11(12):1557.

25. Dean MN, Standaert DG. Levodopa infusion therapies for Parkinson disease. Curr Opin Neurol 2024;37:409–413.

26. de Fabregues O, Falconer D, Heshmat S, et al. Long-Term Evolution of Advanced Parkinson’s Disease Burden: Subgroup Analysis from the 24 Month International PROSPECT Observational Study [abstract]. Mov Disord 2024;39(suppl 1).

